# The impact of COVID-19 on children with oesophageal atresia and/or tracheo-oesophageal fistula (OA/TOF)

**DOI:** 10.1101/2021.02.17.21251622

**Authors:** A. Stewart, C. Smith, S. Eaton, P. De Coppi, J. Wray

## Abstract

**Purpose:** The COVID-19 pandemic has resulted in a global health crisis of unparalleled magnitude. The direct risk to the health of children is low. However, disease containment measures have society-wide impacts. This study explored the pandemic experiences of parents of children with oesophageal atresia/tracheo-oesophageal fistula in the UK.

**Design:** An online forum was conducted using a private group on Facebook, in collaboration with a patient support group. Thematic analysis was used to identify key themes.

**Results:** The online forum ran between 7th November-18th December 2020 with 109 participants. Themes related to healthcare and non-healthcare impacts. Parents experienced changes and limitations to healthcare access, anxiety regarding health risks, “collateral” damage to well-being because of isolation and an impact on finances and employment. Parents described a transition from worry about direct health risks to concern about the impact of isolation on socialisation and development. A process of risk-benefit analysis led some to transition to a more “normal life”, while others continued to isolate. Benefits to their child’s health from isolation, positive experiences with remote healthcare and a gradual easing of anxiety were also identified.

**Implications and relevance:** This study highlights the wide-ranging impact of the COVID-19 pandemic on children and their families. Although focussed on oesophageal atresia/tracheo-oesophageal fistula, the emerging themes will be relevant to many children with complex, chronic health conditions. There are implications for healthcare delivery, including telehealth, during and after the pandemic period. Accurate and consistent messaging is required. Third sector organisations are ideally positioned support this.

## INTRODUCTION

The emergence of SARS-CoV-2 brought about the largest global health crisis for a generation. It is now understood that the direct risk to children from the virus is low, particularly those under 11 years of age.^1 2^ Early data indicated that “high risk” groups for severe disease existed^3^ and as a result individuals, including children, deemed “extremely clinically vulnerable” were advised to “shield”, avoiding contact with others.

One group of vulnerable children are those born with oesophageal atresia/tracheo-oesophageal fistula (OA/TOF). A rare, congenital abnormality, OA/TOF occurs in approximately 1/3500 live births in the UK. While survival rates following surgical repair are excellent, many children experience long-term health challenges.^4^ Swallow dysfunction results in feeding difficulties,^5 6^ oesophageal stricture is common, particularly in the first year of life, and recurrent respiratory infections and chronic cough are evident in 40-52% of adults and children.^5 7^ Hospital readmission with respiratory or gastroenterological issues in the pre-school years is common.^8^

Despite vulnerability to respiratory infection, children with OA/TOF have not experienced severe COVID-19 symptoms.^9^ However, disease containment continues to involve restrictions to social contact, education and non-essential business and has impacted society beyond the immediate risk to health.^10^ Our aim was to understand the wider impact of COVID-19 on children with OA/TOF and their families, learn from their experiences and make recommendations for optimising care for this, and other, rare diseases.

## METHOD

A phenomenological approach underpinned use of an online forum to explore parental experiences of accessing healthcare, education and work, managing health risks and social isolation in the UK^11^ during the COVID-19 pandemic.

Data were collected using a previously described online forum method.^12^ In collaboration with TOFS, the UK support group for OA/TOF, a research-specific, private Facebook group was launched. An experienced member of the TOFS Facebook group, independent of the research team, moderated the forum. This online forum was part of a larger study that was granted ethical approval (20/LO/0098).

### Participants

Convenience sampling was used to recruit parents of children aged 0-18 years with OA/TOF living in the UK. The TOFS support group advertised participation to their members by email and on their Facebook group. Interested parents were asked to apply to join the research Facebook group, with access granted by the moderator after participants agreed to the group “rules”; acknowledging responses would be anonymised and passed to the research team. Participants provided demographic data via a Survey Monkey questionnaire. These data were used to describe group characteristics but were not linked to individual responses.

### Data collection

Six questions were generated by the multi-disciplinary research team and PPI group (including 4 parents of children with OA/TOF), and posted individually by the moderator. A new question was posted once there were no further comments to the existing question. The moderator answered participant questions and prompted for clarification or explanation if required. Participants were also able to respond privately to the moderator. To diversify participation, parents not using Facebook, or not wishing to share information on the forum, could participate via email.

### Data analysis

All responses were anonymised by the moderator. Thematic analysis was conducted^13^ Data were independently coded by 3 members of the research team (AS, JW, CS), followed by group discussion to agree themes. Two thematic maps were generated. Tables of codes with supporting quotes and the maps were reviewed by the PPI group and 2 other members of the research team (PDC, SE) providing data validation from different professional and personal perspectives.

## RESULTS

The online forum ran from 7^th^ November - 18^th^ December 2020. There were 109 members, of whom 65 completed the demographic survey and responded to at least one question. An additional six participants responded by email. Results are presented in Table 1.

**Table 1.** Demographic data

|  |  |  |
| --- | --- | --- |
| Relationship to the child | Mother | 58 (89%) |
|  | Father | 3 (5%) |
|  | Adult with OA/TOF | 1 (2%) |
|  | Did not respond | 3 (5%) |
| Ethnicity | White | 61 (94%) |
|  | Asian or British Asian | 1 (2%) |
|  | Did not respond | 3 (5%) |
| Geographical location | England | 55 (82%) |
|  | Scotland | 10 (15%) |
|  | Wales | 2 (3%) |
| Age of the child | Under 2 years of age | 25 (38%) |
|  | 2-4 years of age | 33 (34%) |
|  | 5-11 years of age | 12 (18%) |
|  | Over 12 years of age | 3 (5%) |
| Type of OA/TOF | OA and TOF repaired within a week of birth | 55 (85%) |
|  | OA and TOF repaired more than a week after birth | 5 (7%) |
|  | OA only | 3 (5%) |
|  | TOF only | 2 (3%) |

### Experiences of accessing healthcare

Participants’ experiences of accessing healthcare are summarised in Figure 1. Further illustrative quotes are shown in Table 2. Themes were grouped into those relating to remote healthcare, delays and cancellations and hospital care. Facilitative factors are highlighted in green boxes, barriers in red.

**Table 2.**
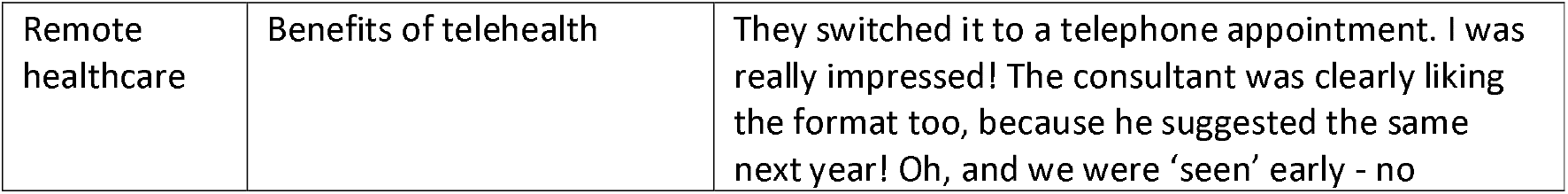
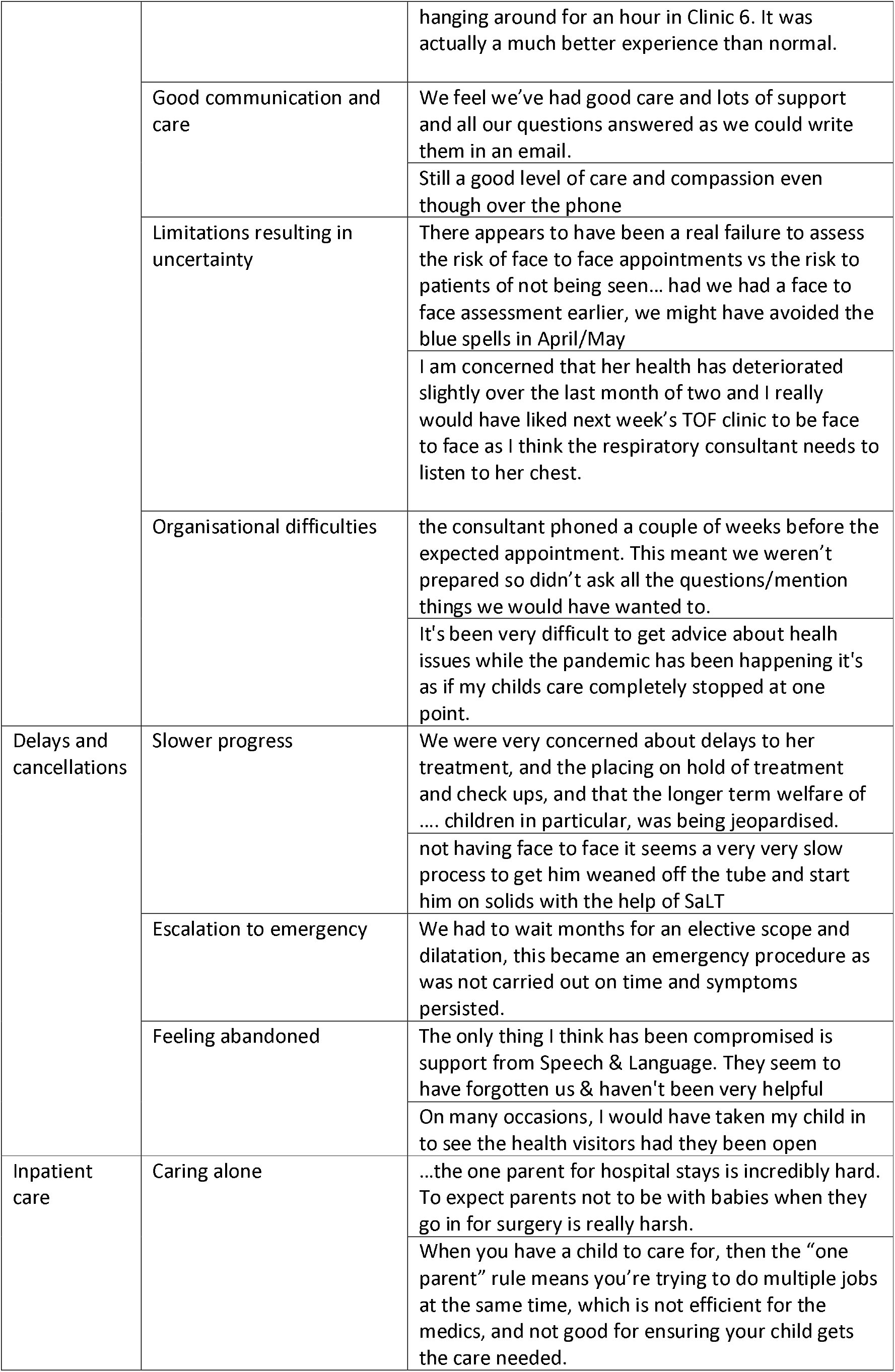
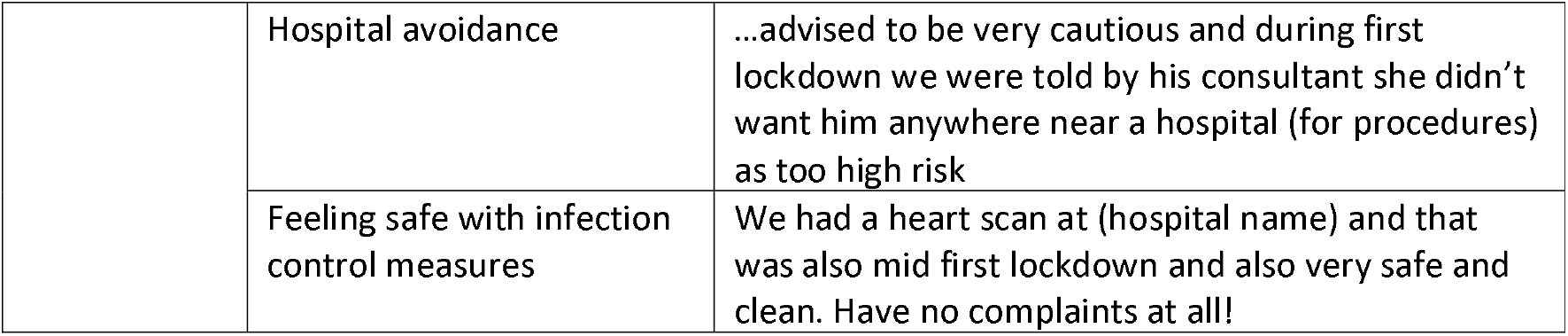
Further illustrative quotes relating to healthcare themes.

**Figure 1.**
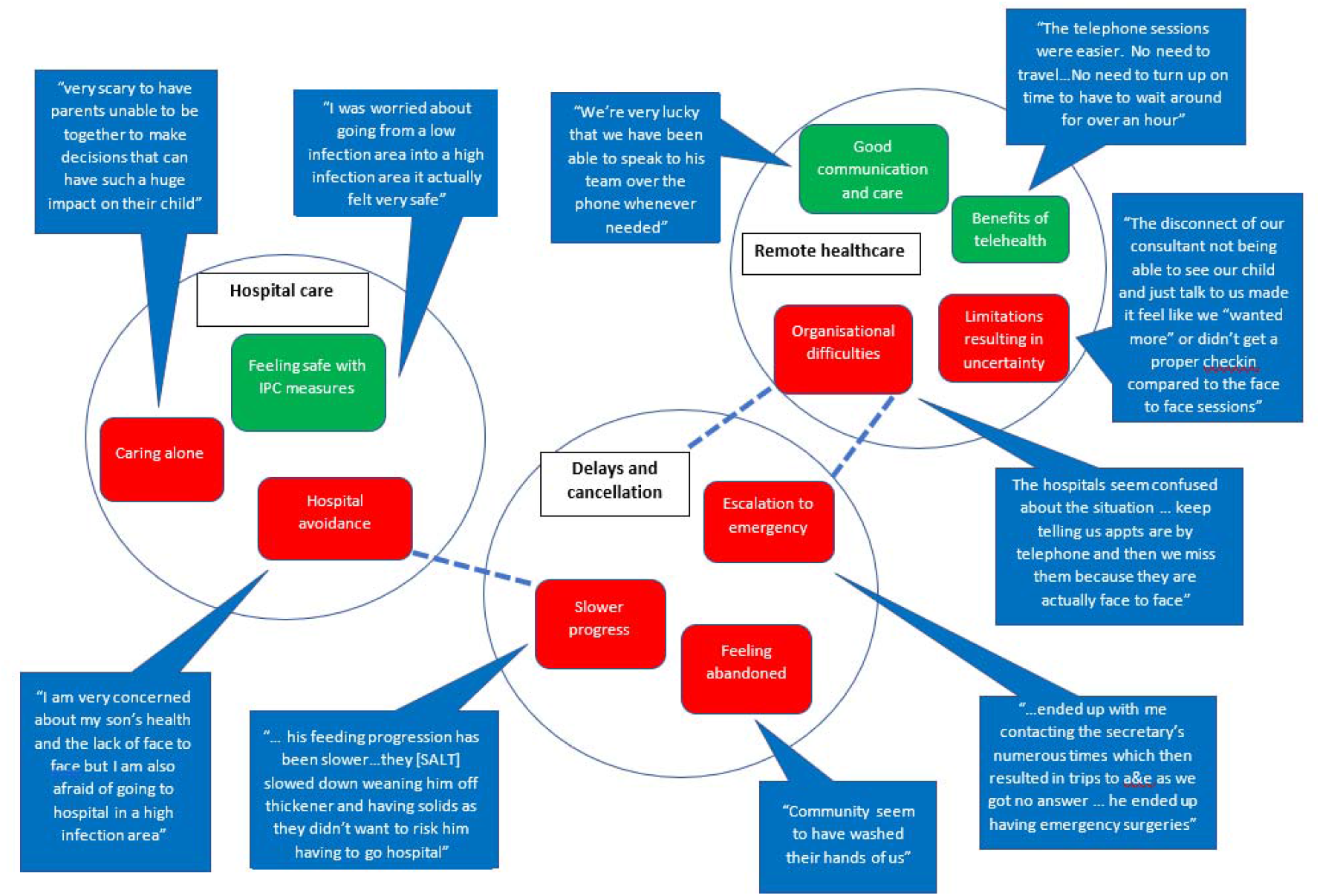
Thematic map with illustrative quotes: healthcare impact

#### Remote healthcare

Access to healthcare changed, with a shift to telehealth (telephone or video) appointments reported by most participants.

Many participants highlighted the ease and convenience of telehealth; no waiting times or travel, not needing to take time off work and not attending busy waiting rooms were benefits. Some parents experienced improved communication with healthcare professionals, having telephone or email contact that was not previously available. Where parents felt connected, remote healthcare was positively received. The value of specialist nurses in achieving good communication was evident.

*“we’ve had huge support from our CNS team who has been amazing throughout the entire journey”*

However, limitations of telehealth were also described. Some parents raised concerns that their child’s health or development was being compromised. Parents reported feeling disconnected from their healthcare team, both as a result of communication or organisational challenges and the limitations of telehealth appointments. A few expressed concern at how their child would cope with face-to-face appointments after a period of not attending in person.

*“I’m worried that my daughter will become withdrawn, nervous, anxious for future appointments”*

#### Delays and cancellations

Delays and cancellations to inpatient and outpatient care were widely reported. Most cancelled appointments had been rebooked.

Participants described concern at anticipated and realised difficulty accessing timely and appropriate care, with stark accounts of feeling abandoned by healthcare services. This was most evident for community services, with Speech and Language Therapy services most frequently cited.

Parents felt that delays impacted directly on their child’s health, including escalation of care to an emergency, and impacts on development and growth.

#### Hospital care

Worries about infection risk and strategies to avoid hospital admission were voiced but all accounts of hospital treatment were positive. Parents felt safe and confident with infection control measures.

The one-parent policy caused the greatest challenge. Participants expressed distress at making decisions regarding surgery and care alone, the absent parent, often the father, being omitted from care, and the challenge of processing information while simultaneously caring for the child.

One parent highlighted the impact of mask-wearing on bonding. Practical challenges, such as not having a parent kitchen and car-parking were also reported. Overall, access to hospital care was reported more positively than community care.

### Wider impact of the pandemic

Themes relating to the wider impact of the pandemic are outlined in Figure 2, with further illustrative quotes provided in Table 3.

**Table 3.** Further illustrative quotes relating to non-healthcare themes

|  |  |
| --- | --- |
| Fear of risk of the child | In the beginning we were extremely concerned and worried about our son catching the virus as months before we had been in hospital for just a cold. |
|  | At the beginning, I think like most people, seeing people on ventilators with respiratory issues was extra concerning for TOFs. |
| Cutting all contact with others | It was extremely stressful, we completely cut off contact with friends and family and shielded, which was difficult and upsetting. |
|  | We completely shielded too to be safe so none of us at home left the house (apart from me walking the dog) March to August |
| Transition to worry about socialisation/development | Now my main worry is him getting the care and support he needs to develop during the crisis. |
| Risk vs benefit | My son is 3 and I personally think he needs to social interaction with other kids and family members. It's a risk- but keeping him cooped up is not natural |
| Feeling reassured | I was relieved when his respiratory consultant explained he no longer needed to shield |
|  | TOFS was ever a great source of support and information by direct posts of latest information and from other parents and their experiences. My son's two consultants were very helpful in putting my mind at rest too. |
| Moving out of isolation | Now we are back to normal and he is going to nursery we are as careful as possible in terms of hygiene but are living as normal now |
|  | It was a worry with our little boy starting Reception... It did ease our minds a little to know the school was doing everything they could. |
| Ongoing fear and isolation | I'm not sure if he should be in school or not but I'm keeping him home. |
|  | We weren't advised to shield but did anyway until August and remain cautious. Our TOF is 10 months now - his grandparents have only held him twice and he has yet to meet Uncles and cousins. |
| Parental mental health impact | I had finally (after poor care at the outset meant we were never seen by the counsellor) started getting help for PTSD. |
|  | For the last six weeks I have been signed off because the pressure overwhelmed me. I have been signed off for a further month and referred for counseling by HR. |
| Impact on work and work-life balance | my husband's ability to work was affected since he was at home with a screaming child usually with him (I was entertaining the 2 year old). |
|  | More work, less time, more pressure to do housework and make the meals due to being at home - and having to create time to make sure we were getting lots of exercise- all his health needs are managed by me. |

**Figure 2.**
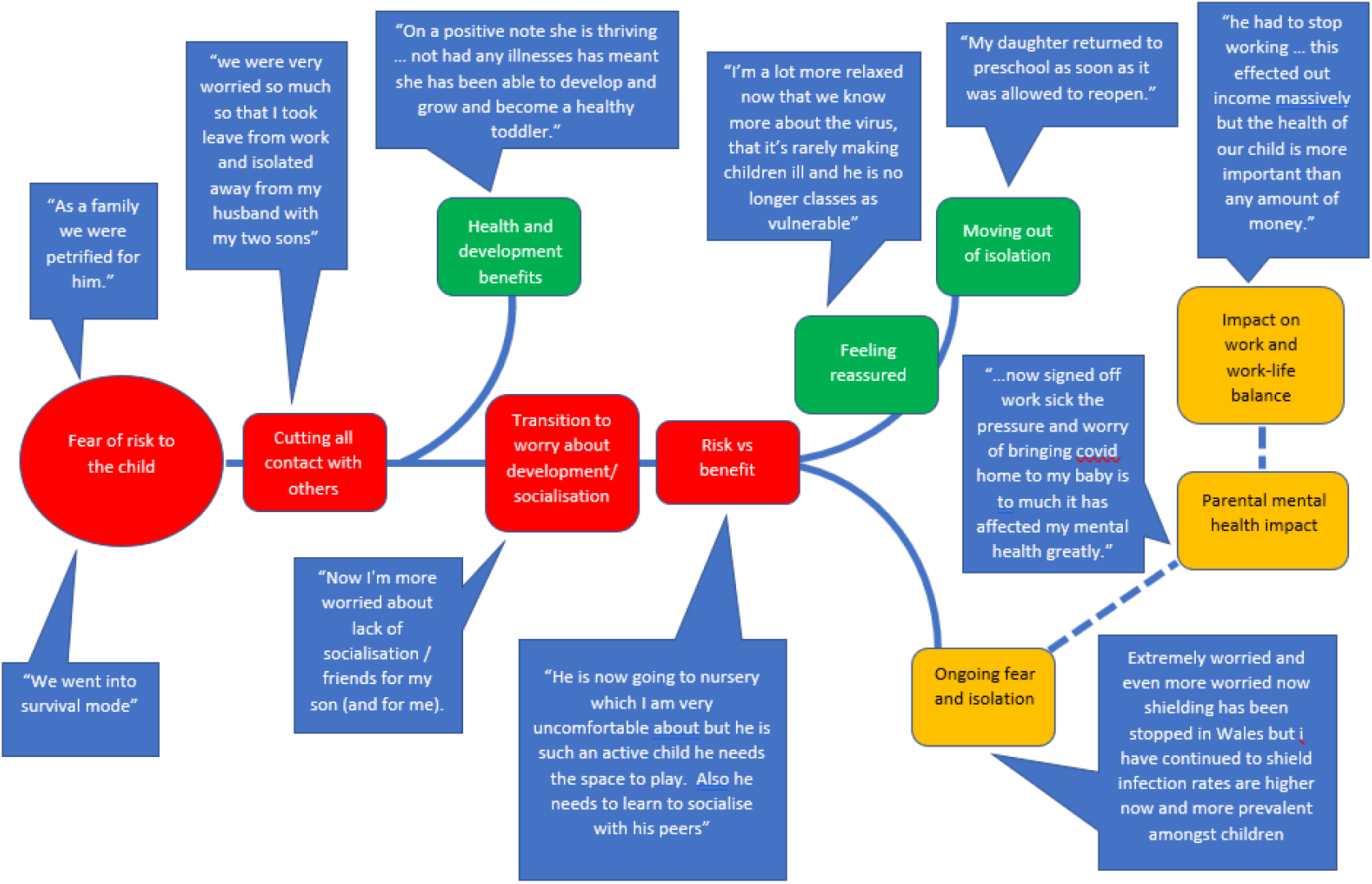
Thematic map: wider impact of COVID-19

#### Fear of risk to the child and cutting contact

Fears for their child’s health were expressed almost universally. Several parents described an overwhelming fear that their child was going to die.

*“I couldn’t shift the feeling again that we that we were going to lose him”*

This led to most participants cutting all contact with others outside their household; some following UK Government advice to “shield”, others without this advice. Whether and when children were advised to shield varied. Some received instruction at the start of the pandemic, some after a couple of months and some not at all. Where shielding advice was not immediate many participants sought information from healthcare teams and the TOFS support group. Participants identified the TOFS website and online peer support as particularly helpful.

*“The TOFS Facebook group is the only place where I have seen useful information about our TOFS”*

Shielding experiences were varied. Some felt gratitude for the family time. For others, balancing work with children at home or the social isolation resulted in high levels of stress.

#### Transitioning to new worries

Many parents transitioned from worrying about their child’s health to worrying about socialisation and development. They described balancing the health benefits of isolation with the risks to well-being.

Participants highlighted the challenge of assessing the risks/benefits of school or childcare attendance. Good communication with the school/nursery and trust in the infection control procedures facilitated attendance. Some reported that infection control measures prevented in-person staff training, disrupting transition to school/nursery. One child was unable to access longer hours in nursery as the parent was unable to go on site to gastrostomy-feed her child. Two parents highlighted to school staff their child’s chronic OA/TOF-related cough to differentiate it from an infectious, COVID-19 related cough.

*“*…*wanted to explain that her TOF cough was normal for her*…*It wasn’t ideal having to try to explain at a distance at the door, but I didn’t want her new teacher to be alarmed (or other kids in the class) when she coughed*.*”*

#### Moving out of isolation and ongoing fear and isolation

Fear led to complete isolation for almost all participants initially, but as the pandemic progressed experiences diverged. For some, increased knowledge provided reassurance. This, coupled with increasing concern about the social and emotional effects of isolation, prompted transition to a more “normal life”, albeit within society-wide restrictions. For others, increased information and knowledge did not provide reassurance, resulting in continued isolation and anxiety, which included continuing with home learning.

#### Health and development benefits

Strikingly, many parents reported benefits of social isolation, highlighting reduced illness and hospitalisations, improved weight gain and improvements to general development.

#### Impact on parental mental health and work-life balance

Parents made direct reference to the impact on their own mental health, with a small number requiring professional support for anxiety or post-traumatic stress disorder.

Combining childcare with home working, managing with limited space and an increased burden of domestic tasks were stress-inducing. Financial hardship was reported. The ability to work was affected by the need to look after children, avoidance of social contact to keep their child safe and parental mental health difficulties.

## DISCUSSION

The online forum allowed for timely gathering of parental insights into the impact of the COVID-19 pandemic on children with OA/TOF. The method engaged a large number of parents from a rare disease cohort, avoided face-to-face contact and minimised burden by allowing for asynchronous participation.

Access to healthcare during the pandemic has been shaped by infection prevention and control measures, limiting face-to-face contact, and prioritisation of “essential” services. This study highlights the significant impact these changes have had.

Telehealth was widely and rapidly adopted. No parents reported having accessed telehealth prior to the pandemic. Benefits, including access to specialist services from geographically distant locations and reduced costs and time to patients reported in this study, have long been recognised.^14^ Recent technological advances in mobile communication, software and high-speed internet have increased feasibility of telehealth ^14^ and effective use in paediatric surgical conditions has been demonstrated.^15 16^

While many parents felt that they received good care remotely, expressing a desire for this to continue post-pandemic, some felt disconnected from their healthcare team and that care was suboptimal. These views are echoed by clinicians.^17^ Our study design prevented linking demographic data to individual responses. However, diagnostic complexity and the age of the child varied within the group. Younger children with OA/TOF tend to have more challenging health needs and post-operative morbidity is greater for some.^8 18^ We propose that satisfaction with telehealth was greater for those at a stable point in their care than those with specific concerns. Dissatisfaction was not reported in other, pre-pandemic studies evaluating use of telehealth with paediatric surgical patients.^15 19^ This may be due to the speed with which telehealth was rolled out, with blanket, rather than targeted, use. It is likely that use of telehealth will continue post-pandemic. The needs of the individual and family must be considered to ensure appropriate use. Providers must address the training needs of the current and future workforce to optimise remote examination and develop effective communication.

Prioritisation of care and redeployment of staff seemingly impacted most on community services. Access to community Speech and Language Therapy appeared particularly poor. The Royal College of Speech and Language Therapists has highlighted the negative impact of redeployment on waiting lists and access to care.^20^ Recent increases in COVID-19 related hospital admissions, the impact of “long-COVID” and staff sickness all continue to threaten access to services. This study highlights the impact that a lack of community services has not only on children’s physical health and development but also on parental mental health. Innovative solutions are required to protect non-acute services in periods of extreme resource demand to minimise “collateral” damage.

The wider harms caused by society wide “lockdown” have been well summarised^10^ and are reflected in our findings; fear and anxiety, displaced non-COVID care, social isolation, stress and loss of income were all reported. Similar themes emerged from research exploring the experiences of parents caring for children with cancer.^21^ Our findings highlight the diversity of experiences, typified by divergent management of anxiety that enabled some to transition to a more normal life while others continued to isolate. Although individual differences in risk evaluation are inevitable, clear communication of the best available evidence would mitigate unnecessary isolation.

Parents highlighted the challenge of obtaining information about the risk to their child. Not all children were identified as “extremely clinically vulnerable” and advice to shield was mixed. Mixed messages and the need for parents to make their own decisions increased anxiety, also identified in parents of children with cancer.^21^ Support groups play an important role in provision of disease-specific information. While individualised assessment may be required, particularly for conditions where morbidity varies, communication between the Government and/or healthcare providers and the charitable sector should be leveraged as a means of sharing accurate and consistent information.

Many parents described the positive impact that isolation had on their child’s health and growth. Exposure to common viruses and other infections is usually an inescapable part of childhood and supports development of a well-functioning immune system^22^ but can necessitate hospital treatment for vulnerable children. The long-term impact of not being exposed is unknown. However, clinicians should be aware of the potential challenge that some parents will face in supporting normal childhood activity with the knowledge that avoidance may improve health post-pandemic.

Chronic cough is common in children with OA/TOF.^7 23^ Interestingly, difficulty differentiating infective coughing from the child’s usual cough was not commonly reported and was not hindering school attendance. TOFS provide excellent resources for families to educate about chronic “TOF cough”, limiting misunderstanding and empowering parents to advocate appropriately.

### Limitations

Despite efforts to facilitate wider involvement, participants were overwhelmingly female, white parents of pre-school children, likely reflecting those most commonly using Facebook and engaging with a support group. We acknowledge that although a wide range of experiences were described, they may not be reflective of the whole OA/TOF community.

Description of group rather than individual demographics supported anonymity but prevented sub-analysis by OA/TOF type or age. Future research should identify whether such factors impact on satisfaction with telehealth and the assessment of risk.

## Conclusion

This study demonstrates the indirect impact that COVID-19 has had on children with a chronic health condition. Parents experienced changes and limitations to healthcare access, anxiety regarding health risks, “collateral” damage to well-being because of isolation and an impact on finances and employment. Benefits to their child’s health from isolation, positive experiences with remote healthcare and a gradual easing of anxiety were also identified.

### What is already known on this topic

- Direct health risk to children from COVID-19 is low but there is a high risk of “collateral” damage from strategies required to contain the virus.
- Patient support groups can be powerful allies in providing accurate and consistent messages, that is particularly useful to those with rare diseases.
- Social media can facilitate rapid and effective data collection in a rare disease cohort.

### What this study adds

- Pandemic-related reduction in healthcare provision has impacted disproportionately on the health and development of children with chronic conditions and on parental mental health.
- Urgent assessment of telehealth policies are required to effectively embed this change to service provision in paediatric practice and ensure appropriate use.
- Isolation has resulted in exceptionally low exposure to usual childhood infections resulting in improved health for children vulnerable to respiratory infection.

## Data Availability

The nature of the qualitative data, whilst anonymised, would potentially allow for identification of participants and the data have therefore not been made available.

## Acknowledgments

The authors wish to thank Joanne Winspear for moderating the Facebook group. Thank you to Diane Stephens and the team at TOFS for their support and guidance in developing the research and facilitating recruitment. Finally, thank you to all the parents who gave their time to share their experiences.

## Funding

Alexandra Stewart is currently undertaking a PhD funded by the National Institute for Health Research (NIHR) (award reference: ICA-CDRF-2018-04-ST2-042).

This publication indicates independent research funded by the National Institute for Health Research (NIHR). The views expressed are those of the authors and not necessarily those of the NHS, the NIHR or the Department of Health and Social Care.

Research at Great Ormond Street Hospital for Children NHS Trust is supported by the NIHR GOS/ICH Biomedical Research Centre.

## Notes

### Competing Interest Statement

The authors have declared no competing interest.

### Author Declarations

This study formed part of a larger study which was granted ethical approval by the London-Central Research Ethics Committee. It was confirmed that specific ethical approval for the online forum was not required as recruitment and data collection was conducted by a charity. Participants had no direct contact with the research team.

## References

1. Ladhani SN, Amin-Chowdhury Z, Davies HG, et al. COVID-19 in children: analysis of the first pandemic peak in England. Arch Dis Child 2020;105(12):1180–85. doi: 10.1136/archdischild-2020-320042 [published Online First: 2020/08/17]

2. European Centre for Disease Prevention and Control. COVID-19 in children and the role of school settings in transmission - first update. Stockholm, 2020.

3. Yang J, Zheng Y, Gou X, et al. Prevalence of comorbidities and its effects in patients infected with SARS-CoV-2: a systematic review and meta-analysis. Int J Infect Dis 2020;94:91–95. doi: 10.1016/j.ijid.2020.03.017 [published Online First: 2020/03/17]

4. van Lennep M, Singendonk MMJ, Dall’Oglio L, et al. Oesophageal atresia. Nat Rev Dis Primers 2019;5(1):26. doi: 10.1038/s41572-019-0077-0 [published Online First: 2019/04/20]

5. Connor MJ, Springford LR, Kapetanakis VV, et al. Esophageal atresia and transitional care--step 1: a systematic review and meta-analysis of the literature to define the prevalence of chronic long-term problems. Am J Surg 2015;209(4):747–59. doi: 10.1016/j.amjsurg.2014.09.019 [published Online First: 2015/01/22]

6. Mahoney L, Rosen R. Feeding Difficulties in Children with Esophageal Atresia. Paediatr Respir Rev 2016;19:21–7. doi: 10.1016/j.prrv.2015.06.002 [published Online First: 2015/07/15]

7. Kovesi T. Aspiration Risk and Respiratory Complications in Patients with Esophageal Atresia. Front Pediatr 2017;5:62. doi: 10.3389/fped.2017.00062 [published Online First: 2017/04/20]

8. Scheider A, Blanc, S., Bonnard, A et al. Results from the French national esophageal atresia register: 1 yr outcome. Orphanet journal of rare diseases 2014;9:206.

9. TOFS. 2021 [Available from: https://www.tofs.org.uk/news/2020/03/coronavirus-update.aspx accessed 28th January 2021.

10. Douglas M, Katikireddi SV, Taulbut M, et al. Mitigating the wider health effects of covid-19 pandemic response. BMJ 2020;369:m1557. doi: 10.1136/bmj.m1557 [published Online First: 2020/04/29]

11. Neubauer BE, Witkop CT, Varpio L. How phenomenology can help us learn from the experiences of others. Perspect Med Educ 2019;8(2):90–97. doi: 10.1007/s40037-019-0509-2 [published Online First: 2019/04/07]

12. Wray J, Brown K, Tregay J, et al. Parents’ Experiences of Caring for Their Child at the Time of Discharge After Cardiac Surgery and During the Postdischarge Period: Qualitative Study Using an Online Forum. Journal of Medical Internet Research 2018;20(5) doi: 10.2196/jmir.9104

13. Braun V, Clarke V. Using thematic analysis in psychology. Qualitative research in psychology 2006;3(2):77–101.

14. Dorsey ER, Topol EJ. State of Telehealth. N Engl J Med 2016;375(2):154–61. doi: 10.1056/NEJMra1601705 [published Online First: 2016/07/15]

15. Dean P, O’Donnell M, Zhou L, et al. Improving value and access to specialty medical care for families: a pediatric surgery telehealth program. Can J Surg 2019;62(6):436–41. doi: 10.1503/cjs.005918 [published Online First: 2019/11/30]

16. Goedeke J, Ertl A, Zoller D, et al. Telemedicine for pediatric surgical outpatient follow-up: A prospective, randomized single-center trial. J Pediatr Surg 2019;54(1):200–07. doi: 10.1016/j.jpedsurg.2018.10.014 [published Online First: 2018/10/23]

17. Kemp MT, Liesman DR, Williams AM, et al. Surgery Provider Perceptions on Telehealth Visits During the COVID-19 Pandemic: Room for Improvement. J Surg Res 2020;260:300–06. doi: 10.1016/j.jss.2020.11.034 [published Online First: 2020/12/29]

18. Svoboda E, Fruithof J, Widenmann-Grolig A, et al. A patient led, international study of long term outcomes of esophageal atresia: EAT 1. Journal of Pediatric Surgery 2018;53(4):610–15.

19. Kohler JE, Falcone RA, Jr., Fallat ME. Rural health, telemedicine and access for pediatric surgery. Curr Opin Pediatr 2019;31(3):391–98. doi: 10.1097/MOP.0000000000000763 [published Online First: 2019/05/16]

20. Royal College of Speech and Language Therapists. Redeployment of staff and impact on outcomes for service. London, 2020:1–5.

21. Darlington AE, Morgan JE, Wagland R, et al. COVID-19 and children with cancer: Parents’ experiences, anxieties and support needs. Pediatr Blood Cancer 2021;68(2):e28790. doi: 10.1002/pbc.28790 [published Online First: 2020/11/22]

22. Ball TM, Castro-Rodriguez JA, Griffith KA, et al. Siblings, day-care attendance, and the risk of asthma and wheezing during childhood. New England journal of medicine 2000;343(8):538–43.

23. Sadreameli SC, McGrath-Morrow SA. Respiratory Care of Infants and Children with Congenital Tracheo-Oesophageal Fistula and Oesophageal Atresia. Paediatr Respir Rev 2016;17:16–23. doi: 10.1016/j.prrv.2015.02.005 [published Online First: 2015/03/25]

